# Research Letter: Application of GPT-4 to select next-step antidepressant treatment in major depression

**DOI:** 10.1101/2023.04.14.23288595

**Authors:** Roy H. Perlis

## Abstract

**Introduction:** Large language models perform well on a range of academic tasks including medical examinations. The performance of this class of models in psychopharmacology has not been explored.

**Method:** Chat GPT-plus, implementing the GPT-4 large language model, was presented with each of 10 previously-studied antidepressant prescribing vignettes in randomized order, with results regenerated 5 times to evaluate stability of responses. Results were compared to expert consensus.

**Results:** At least one of the optimal medication choices was included among the best choices in 38/50 (76%) vignettes: 5/5 for 7 vignettes, 3/5 for 1, and 0/5 for 2. At least one of the poor choice or contraindicated medications was included among the choices considered optimal or good in 24/50 (48%) of vignettes. The model provided as rationale for treatment selection multiple heuristics including avoiding prior unsuccessful medications, avoiding adverse effects based on comorbidities, and generalizing within medication class.

**Conclusion:** The model appeared to identify and apply a number of heuristics commonly applied in psychopharmacologic clinical practice. However, the inclusion of less optimal recommendations indicates that large language models may pose a substantial risk if routinely applied to guide psychopharmacologic treatment without further monitoring.

## Introduction

Large language models have been shown to perform well on a range of academic tasks^1^, with GPT-4 achieving above-median scores on the Medical College Admission Test^2^ and passing the US Medical Licensing Exam^3^. These models have begun to be explored in more focused medical problem-solving as well^4^. One such class of problems is treatment selection in psychiatry, often cited as an opportunity to apply artificial intelligence to better match patients with optimal treatments.

In prior work, the author and colleagues found that many predictors of antidepressant treatment outcome are non-specific^5^, such that machine learning models performed modestly in treatment selection. In routine clinical practice, treatment selection is simple to implement in terms of rules^6^, and we have previously shown that clinicians commonly apply such heuristics in selecting treatments^7^, specifically as they relate to comorbidities and adverse effect profiles. However, this process may become more arbitrary when multiple rules conflict, and different weights must be assigned, or when there remain numerous reasonable options after application of rules.

In the present investigation, the author sought to evaluate the performance of one such model, GPT 4^2^, on a basic psychopharmacologic task, namely selecting the optimal antidepressant treatment after non-response to an initial treatment. The author and colleagues previously created and validated brief vignettes about adults with major depressive disorder illustrating aspects of psychopharmacologic decisionmaking, and presented these vignettes to a panel of experienced clinicians^8^. These clinicians were asked to rank the appropriateness of a list of antidepressants for each vignette. We then presented these vignettes to a group of clinicians via web-based survey, along with a decision support dashboard, as part of an investigation about explainable artificial intelligence^8^. Here, these validated vignettes were applied to investigate the performance of a publicly-available large language model on this task, in order to gain insight about strengths and limitations of this model as it pertains to possible application in psychiatry, and medicine more broadly.

## Method

Chat-GPT-plus, which implements the GPT 4 large language model^2^, was presented with each of the 10 vignettes in randomized order, with random variation in age and distractor medications, with results regenerated 5 times to evaluate stability of responses. All interactions were recorded between April 12, 2023 and April 14, 2023. The chat was re-initialized for each vignette. In order to receive recommendations, a prompt was developed that indicated that the question was not for clinical application; absent such a prompt, the model declined to answer, indicating it was not able to provide medical advice. The prompt sought to mimic the original rating task, presenting a vignette followed by the treatment options. The key element of the prompt was, “Please rate each of the following medications, from “Best choice for this patient” to “Good choice for this patient” to “Less good choice for this patient” to “Bad choice for this patient, or contraindicated”“. Results were then compared to those of expert clinicians in terms of a) whether the optimal expert-curated treatment was identified among the best options, and b) whether a less good or contraindicated treatment was identified among the best or good options, with descriptive statistics (proportion of correct trials) calculated. The author then manually curated model-provided explanations in order to gain further insight into the processes underlying treatment selection.

## Results

With the standard prompt, at least one of the optimal medication choices was included among the best choices in 38/50 (76%) (5/5 for 7 vignettes, 3/5 for 1, and 0/5 for 2). At least one of the poor choice or contraindicated medications was included among the best or good choices in 24/50 (48%) (including 2 vignettes with 5/5, 2 with 4/5, one with 3/5, 3 with 1/5, one with 0/5).

A number of phenomena were apparent in the narrative explanations for treatment choice (Supplemental Materials). Prior treatment non-response was recognized as a reason not to re-use the same medication. However, the model did not consistently generalize across mechanistic classes – for example, recommending another SSRI after initial SSRI failure – although it did in some cases, noting an SSRI “may not be the best choice for this patient, as they have not worked for her before.” The model also recognized escitalopram as an enantiomer of citalopram.

The concept of indication was recognized (i.e., noting that a treatment was used for depression, while avoiding milnacipran as not approved for depression, and down-rating other antidepressants as having less evidence for efficacy in some contexts), including other indications such as OCD and diabetic neuropathy. In one case, however, an indication for bupropion (smoking cessation) was identified as a rationale for scoring that medication as less favorable.

Potential adverse effects that may exacerbate medical comorbidities were recognized as reasons to avoid medications. For example, among individuals with a history of arrhythmia, tricyclic antidepressants were avoided (although not QT-prolonging SSRIs), as was lowering seizure threshold in individuals with prior seizures – although on one run the model identified lower seizure threshold as a reason for caution but still listed bupropion as a good treatment option.

However, the tendency to cause a given AE was apparently hallucinated in some cases, such as avoiding anticholinergic medications in atopic dermatitis or bupropion in atrial fibrillation.

In some cases, the model correctly matched adverse effect profile to patient symptoms or comorbidities, most notably with obesity (favoring bupropion, e.g.) and weight loss (favoring mirtazapine). For example, it noted “This medication may be particularly beneficial … because it can help with both depression and fatigue, and it has a lower risk of weight gain compared to some other antidepressants.” However, while recognizing weight gain as a side effect (“it has a higher risk of weight gain, which may not be ideal for a patient with obesity”), it often favored medications with weight gain risk as good options for obese patients.

Another heuristic that emerged in some model runs was the notion of drug-drug interactions. Interactions were largely hallucinated (e.g., SSRIs with amoxicillin, while amoxicillin is not a CYP450 substrate; clarithromycin, a CYP450 3A4 inhibitor, with non CYP3A4 substrates; and essentially any medication with MAO inhibitors). The model also indicated a preference for newer drugs on the basis of overall tolerability and risk for drug interactions; in nearly all cases, tricyclic antidepressants and MAOIs were disfavored, noting side effect burden and for the latter, dietary restrictions and (mostly hallucinated) drug-drug interactions. (Notably, the one circumstance a tricyclic antidepressant was favored was a vignette identifying a history of obsessive-compulsive disorder, for which clomipramine was identified as an optimal choice).

## Discussion

A publicly-available large language model, GPT-4, provided generally reasonable medication recommendations for pharmacotherapy of major depression after initial antidepressant nonresponse, consistently identifying optimal treatment in 76% of vignettes, but also advised less optimal or contraindicated approaches for all but one vignette. By comparison, in a prior clinician survey using these vignettes, an optimal treatment was selected for 36% of vignettes presented^8^. The model appeared to identify and apply a number of common heuristics in clinical practice – avoiding medications that had not shown efficacy in the past, favoring better-tolerated medications, avoiding medications with adverse effects that could worsen medical comorbidities, and avoiding drug-drug interactions. However, it sometimes failed to consistently apply a medication contraindication (regarding bupropion and seizures, e.g.), sometimes advised medication associated with weight gain to obese patients, and demonstrated a lack of ability to reliably identify and apply drug-drug interactions.

In prior work, the author and colleagues found evidence that clinicians commonly apply simple heuristics in selecting antidepressant treatment^7^. This study suggests that a large language model also applies such heuristics without their being specified explicitly. As many of these heuristics would likely apply across psychopharmacologic decisionmaking, this represents an encouraging observation for application of these models in prescribing more generally. On the other hand, given the frequency with which adverse effects were hallucinated, or not applied as contraindications, extreme caution is needed in applying any such recommendations.

GPT-4 consistently responded with language cautioning against clinical application; without framing prompts in terms of teaching or some other non-clinical use, it would not return recommendations. (Interestingly, these warnings often noted that GPT-4 is not a ‘licensed’ psychiatrist, despite the reality that most antidepressants are prescribed by non-psychiatrists). Since trivial tweaks to the prompt did enable receipt of medical advice, albeit with caveats, this work also suggests the likelihood that these models will be applied in clinical settings as they become more accessible, regardless of warnings to the contrary. The implication of such clinical use requires further study.

More broadly, this work reinforces the importance of developing and applying systematic measures of quality in psychopharmacology, in order to reliably understand the impact of technology deployment. There is growing awareness of the extent to which machine learning models may be widely deployed despite poor performance^9^ and biased outcomes^10^. The ease with which large language models may be applied to guide prescribing, and the superficially reasonable results we identify, indicates the need to track when such algorithms are applied and how they impact outcomes.

## Supporting information

Supplemental Methods and Results

## Data Availability

Results available in supplemental materials

## Acknowledgments

Dr. Perlis is supported by the National Institute of Mental Health (R01MH116270)

