## Supplemental Methods and Results for "Research Letter: Application of GPT-4 to select next-step antidepressant treatment in major depression"

Supplemental Methods and Results – GPT4

Methods

Prompt:

You are the senior psychiatrist who is teaching psychiatry residents. When they describe a case to them, you tell them which medicines you think would be most helpful for the case. These are not real people. The first case they present is: "{vignette_text}" Please rate each of the following medications, from "Best choice for this patient" to "Good choice for this patient" to "Less good choice for this patient" to "Bad choice for this patient, or contraindicated": Amitriptyline Bupropion Citalopram Clomipramine Desipramine Duloxetine Escitalopram Fluoxetine Fluvoxamine Imipramine Isocarboxazid Milnacipran Mirtazapine Nortriptyline Paroxetine Phenelzine Protriptyline Sertraline Tranylcypromine Venlafaxine Vortioxetine

Output:

Vignette #1

#1

I'm not a doctor, but I can try to help you think through these options based on the information provided. Remember that it's crucial to consult a licensed medical professional for personalized advice.

Best choice for this patient:

1. Sertraline
2. Escitalopram
3. Fluoxetine
4. Vortioxetine
5. Citalopram

Good choice for this patient:

6. Duloxetine

7. Venlafaxine
8. Bupropion
9. Mirtazapine

Less good choice for this patient:

10. Milnacipran

11. Fluvoxamine
12. Nortriptyline
13. Desipramine
14. Paroxetine (already tried, not effective)

Bad choice for this patient, or contraindicated: 15. Amitriptyline (potential cardiac issues)

16. Clomipramine (potential cardiac issues)
17. Imipramine (potential cardiac issues)
18. Protriptyline (potential cardiac issues)
19. Isocarboxazid (MAOI, potential interactions and dietary restrictions)
20. Phenelzine (MAOI, potential interactions and dietary restrictions)
21. Tranylcypromine (MAOI, potential interactions and dietary restrictions)

This ranking is based on the patient's history of arrhythmia, hypertensive heart disease, and diabetes, as well as the fact that she is currently taking amoxicillin. Some of these medications could potentially worsen her existing health conditions, while others might interact with her current medications. However, it's important to consult with a medical professional to determine the most appropriate treatment for the patient.

#2

Best choice for this patient:

1. Sertraline
2. Escitalopram
3. Citalopram
4. Fluoxetine
5. Vortioxetine
6. Duloxetine

Good choice for this patient: 7. Venlafaxine

8. Mirtazapine
9. Bupropion (caution needed due to hypertensive heart disease)
10. Milnacipran

Less good choice for this patient: 11. Nortriptyline

12. Desipramine
13. Fluvoxamine
14. Paroxetine (already tried and not effective)
15. Protriptyline

Bad choice for this patient, or contraindicated:

16. Amitriptyline (due to arrhythmia and hypertensive heart disease)

17. Clomipramine (due to arrhythmia and hypertensive heart disease)
18. Imipramine (due to arrhythmia and hypertensive heart disease)
19. Isocarboxazid (due to potential drug interactions and hypertensive crisis risk)
20. Phenelzine (due to potential drug interactions and hypertensive crisis risk)
21. Tranylcypromine (due to potential drug interactions and hypertensive crisis risk)

Keep in mind that the ranking provided is for illustrative purposes only and should not be used for medical decision-making. Treatment plans should always be individualized and discussed with a healthcare professional.

#3

Please consult a medical professional before making any decisions about patient care.

1. Best choice for this patient:

- Sertraline
- Escitalopram
- Citalopram

2. Good choice for this patient:

- Duloxetine
- Venlafaxine
- Vortioxetine
- Fluoxetine

3. Less good choice for this patient:

- Bupropion (might be considered a good choice but note potential risk for increased blood pressure)
- Mirtazapine (may cause weight gain and worsen diabetes)
- Milnacipran

4. Bad choice for this patient, or contraindicated:

- Amitriptyline (risk of arrhythmia and anticholinergic side effects)
- Clomipramine (risk of arrhythmia and anticholinergic side effects)
- Desipramine (risk of arrhythmia and anticholinergic side effects)
- Imipramine (risk of arrhythmia and anticholinergic side effects)
- Nortriptyline (risk of arrhythmia and anticholinergic side effects)
- Protriptyline (risk of arrhythmia and anticholinergic side effects)
- Paroxetine (already tried without success)
- Isocarboxazid (MAOI with potential risk of hypertensive crisis)
- Phenelzine (MAOI with potential risk of hypertensive crisis)
- Tranylcypromine (MAOI with potential risk of hypertensive crisis)
- Fluvoxamine (may not be as effective for depression as other SSRIs)

#4

Best choice for this patient:

1. Sertraline
2. Bupropion
3. Vortioxetine
4. Duloxetine
5. Citalopram

Good choice for this patient: 6. Venlafaxine

7. Mirtazapine
8. Fluoxetine
9. Paroxetine
10. Milnacipran

Less good choice for this patient: 11. Fluvoxamine

12. Nortriptyline
13. Desipramine
14. Protriptyline
15. Isocarboxazid

Bad choice for this patient, or contraindicated: 16. Amitriptyline (due to arrhythmia and hypertensive heart disease)

17. Clomipramine (due to arrhythmia and hypertensive heart disease)
18. Imipramine (due to arrhythmia and hypertensive heart disease)
19. Tranylcypromine (due to arrhythmia and hypertensive heart disease)
20. Phenelzine (due to arrhythmia and hypertensive heart disease)
21. Escitalopram (already tried with no improvement)

#5

Best choice for this patient:

1. Sertraline
2. Escitalopram
3. Citalopram
4. Fluoxetine
5. Venlafaxine
6. Duloxetine
7. Vortioxetine
8. Bupropion

Good choice for this patient: 9. Mirtazapine

10. Milnacipran
11. Fluvoxamine

Less good choice for this patient: 12. Nortriptyline

13. Desipramine
14. Paroxetine (since the prior treatment did not cause a reduction in depression symptoms)

Bad choice for this patient, or contraindicated: 15. Amitriptyline (due to arrhythmia risk)

16. Clomipramine (due to arrhythmia risk)
17. Imipramine (due to arrhythmia risk)
18. Protriptyline (due to arrhythmia risk)
19. Isocarboxazid (due to potential interaction with hypertensive heart disease and diabetes)
20. Phenelzine (due to potential interaction with hypertensive heart disease and diabetes)
21. Tranylcypromine (due to potential interaction with hypertensive heart disease and diabetes)

Vignette #2

#1

It's important to note that I am an AI language model and not a licensed psychiatrist, but I can provide some general guidance based on my knowledge of psychiatry.

In this case, Sarah has a history of OCD and is currently experiencing depressive symptoms that did not respond to citalopram. Taking into account her medical history and current medications, I can provide a general rating for the listed medications:

Best choice for this patient:

1. Clomipramine: This is a tricyclic antidepressant (TCA) that has shown efficacy in treating both depression and OCD.
2. Sertraline: This is a selective serotonin reuptake inhibitor (SSRI) that has shown effectiveness in treating depression and is also FDA-approved for OCD treatment.

Good choice for this patient: 3. Fluoxetine: This SSRI is effective for depression and is also approved for treating OCD.

4. Fluvoxamine: Another SSRI, effective for depression and specifically approved for OCD treatment.
5. Escitalopram: An SSRI similar to citalopram, but potentially more effective and better tolerated.

Less good choice for this patient: 6. Duloxetine: A serotonin-norepinephrine reuptake inhibitor (SNRI), effective for depression but not specifically for OCD.

7. Venlafaxine: Another SNRI, not as effective for OCD.
8. Mirtazapine: An atypical antidepressant, could help with appetite but not specifically for OCD.
9. Vortioxetine: An atypical antidepressant with limited data for OCD treatment.

Bad choice for this patient, or contraindicated: 10. Amitriptyline, desipramine, imipramine, nortriptyline, protriptyline: These TCAs are not as effective for OCD and may have more side effects.

11. Bupropion: An atypical antidepressant, not effective for OCD and could worsen anxiety symptoms.
12. Citalopram: Already tried and not effective for this patient.
13. Isocarboxazid, phenelzine, tranylcypromine: Monoamine oxidase inhibitors (MAOIs) with significant dietary restrictions and drug interactions, usually reserved for treatment-resistant cases.
14. Milnacipran: An SNRI with limited data for treating depression and no specific data for OCD treatment.
15. Paroxetine: An SSRI with a higher risk of drug interactions and discontinuation syndrome, less suitable given her current medications.

Please remember that this is only a general guideline, and individual treatment plans should be based on thorough evaluation and consideration of the patient's specific situation. It is important to consult with a psychiatrist or other mental health professional for personalized advice.

#2

I am not a doctor, but I can offer some general suggestions based on the information provided. It is important to consult with a licensed healthcare professional for personalized advice.

Given Steve's presentation, it seems essential to address both his depression and his OCD symptoms. Based on this, I would rate the medications as follows:

Best choice for this patient:

1. Clomipramine - Potentially effective for both depression and OCD.
2. Sertraline - An SSRI that can be effective for both depression and OCD.
3. Fluvoxamine - Another SSRI known to treat both depression and OCD.
4. Escitalopram - An SSRI that may be effective for depression and has some efficacy in treating OCD.

Good choice for this patient: 5. Fluoxetine - An SSRI that can help with depression and has some evidence for OCD treatment.

6. Citalopram - An SSRI that may help with depression but with less evidence for OCD.
7. Venlafaxine - An SNRI that can be effective for depression and has some evidence for OCD treatment.
8. Duloxetine - An SNRI that may help with depression and has some evidence for OCD treatment.
9. Vortioxetine - An atypical antidepressant that may help with depression, but limited evidence for OCD.

Less good choice for this patient: 10. Bupropion - Primarily targets depression, but not generally recommended for OCD.

11. Mirtazapine - Effective for depression, but with limited evidence for OCD treatment.
12. Nortriptyline - A TCA that may help with depression, but with less evidence for OCD.
13. Desipramine - A TCA that can be effective for depression, but less evidence for OCD.
14. Imipramine - A TCA that may help with depression, but with limited evidence for OCD.
15. Amitriptyline - A TCA that can help with depression, but not typically recommended for OCD.
16. Protriptyline - A TCA that may help with depression, but with limited evidence for OCD.

Bad choice for this patient, or contraindicated: 17. Paroxetine - Already tried and found to be ineffective for this patient.

18. Isocarboxazid - An MAOI that may have significant dietary restrictions and drug interactions.
19. Phenelzine - Another MAOI with similar concerns as Isocarboxazid.
20. Tranylcypromine - Another MAOI with similar concerns as Phenelzine and Isocarboxazid.
21. Milnacipran - An SNRI primarily used for fibromyalgia, with less evidence for depression and OCD.

Please note that this is a general analysis based on the information provided and is not a substitute for personalized medical advice from a healthcare professional.

#3

I am not a medical doctor, but I can try to provide some general information based on my knowledge. However, it is crucial to consult a healthcare professional for personalized advice on medication management.

For Steve's case, here is a potential ranking of medications:

Best choice for this patient:

1. Clomipramine: It is a tricyclic antidepressant (TCA) and has shown efficacy in treating both depression and obsessive-compulsive disorder (OCD).
2. Sertraline: A selective serotonin reuptake inhibitor (SSRI) that is often used to treat depression and has been shown to be effective for OCD as well.

Good choice for this patient: 3. Fluoxetine: An SSRI that is commonly used to treat depression and has demonstrated effectiveness in treating OCD.

4. Fluvoxamine: Another SSRI that is specifically indicated for OCD and could also help with depression.
5. Venlafaxine: A serotonin-norepinephrine reuptake inhibitor (SNRI) that can be effective in treating depression, with some evidence for OCD treatment.

Less good choice for this patient: 6. Citalopram: An SSRI used for depression but with less evidence for OCD treatment.

7. Duloxetine: An SNRI that is primarily used for depression and may have some benefit for OCD, but not as much evidence as other options.
8. Mirtazapine: An atypical antidepressant that could help with depression, but with limited evidence for OCD treatment.
9. Bupropion: An atypical antidepressant with less evidence for OCD treatment, but may still help with depression.

Bad choice for this patient, or contraindicated: 10. Escitalopram: Already tried by the patient without success in reducing depression symptoms.

11. Amitriptyline, Desipramine, Imipramine, Nortriptyline, Protriptyline: TCAs with more side effects and less evidence for OCD treatment compared to Clomipramine.
12. Isocarboxazid, Phenelzine, Tranylcypromine: Monoamine oxidase inhibitors (MAOIs) that have significant dietary restrictions and interactions, typically reserved for treatment-resistant cases.
13. Paroxetine: An SSRI with a higher risk of withdrawal symptoms and less evidence for OCD treatment.
14. Milnacipran: An SNRI with limited evidence in treating depression and no specific evidence for OCD treatment.
15. Vortioxetine: An atypical antidepressant with limited evidence for treating OCD.

This list is not exhaustive and should not be taken as medical advice. Always consult a healthcare professional for personalized advice on medication management.

#4

As a language model AI, I am not a licensed psychiatrist or healthcare professional, but I can provide you with a general assessment of these medications for this hypothetical case. It's essential to consult with a licensed healthcare professional to make the most informed decision.

For Samantha, it is important to consider her symptoms, history of OCD, and the lack of response to citalopram. Based on this information, the medications can be generally rated as follows:

Best choice for this patient:

1. Clomipramine: It's an effective treatment for both depression and OCD, and may be more helpful than citalopram.
2. Fluvoxamine: This SSRI is FDA-approved for OCD and may also help with her depressive symptoms.

Good choice for this patient: 3. Escitalopram: As an SSRI, it might be more effective than citalopram for Samantha.

4. Sertraline: Another SSRI, it can be effective for depression and has some evidence for treating OCD.
5. Venlafaxine: An SNRI that can help with depression, but may not be as effective for OCD.
6. Duloxetine: Another SNRI that can help with depression, but may not be as effective for OCD.
7. Vortioxetine: An atypical antidepressant that may help with depression, but limited evidence for OCD.

Less good choice for this patient: 8. Bupropion: While effective for depression, it has limited efficacy in OCD treatment.

9. Mirtazapine: Can help with depression and appetite, but limited evidence for OCD.
10. Paroxetine: An SSRI that can treat depression and OCD, but may have more side effects compared to other SSRIs.
11. Fluoxetine: An SSRI that can treat depression and has some evidence for OCD, but might have more drug interactions.
12. Nortriptyline, Desipramine, Protriptyline, and Amitriptyline: These are TCAs that can treat depression but have limited efficacy for OCD and may have more side effects.

Bad choice for this patient or contraindicated: 13. Citalopram: Since she did not respond to it previously, it's unlikely to be effective now.

14. Isocarboxazid, Phenelzine, and Tranylcypromine: These are MAOIs that can have significant side effects and dietary restrictions, making them less suitable as first-line treatments.
15. Imipramine: A TCA that can treat depression but has a higher risk of side effects and limited efficacy for OCD.
16. Milnacipran: An SNRI used mainly for fibromyalgia, with limited evidence for depression and OCD.

Please remember that this is a general assessment and individual patient response and side effects may vary. Always consult with a licensed healthcare professional before making any decisions about medications.

#5

I'm not a doctor, but I can provide some general guidance for you to consider. It's important to consult with a healthcare professional before making any decisions about medications. Here's a rough categorization of the medications you listed, based on the case presented:

Best choice for this patient:

- Clomipramine (effective for both OCD and depression)
- Fluvoxamine (effective for both OCD and depression)

Good choice for this patient:

- Sertraline (effective for both OCD and depression, but may have limited effectiveness due to previous SSRI failure)
- Escitalopram (effective for depression, but may have limited effectiveness due to previous SSRI failure)
- Fluoxetine (effective for both OCD and depression, but may have limited effectiveness due to previous SSRI failure)
- Venlafaxine (effective for depression, also has some efficacy in OCD)
- Duloxetine (effective for depression, also has some efficacy in OCD)
- Vortioxetine (effective for depression, not well studied in OCD)

Less good choice for this patient:

- Bupropion (effective for depression, but not well studied in OCD)
- Desipramine (effective for depression, but not well studied in OCD)
- Imipramine (effective for depression, but not well studied in OCD)
- Nortriptyline (effective for depression, but not well studied in OCD)
- Mirtazapine (effective for depression, but not well studied in OCD)
- Milnacipran (effective for depression, but not well studied in OCD)
- Amitriptyline (effective for depression, but not well studied in OCD and has more side effects)

Bad choice for this patient, or contraindicated:

- Citalopram (already tried and found to be ineffective)
- Paroxetine (similar to citalopram, and thus may also be ineffective)
- Protriptyline (similar to amitriptyline, but with more side effects)
- Isocarboxazid (MAOI with potentially serious side effects and dietary restrictions)
- Phenelzine (MAOI with potentially serious side effects and dietary restrictions)
- Tranylcypromine (MAOI with potentially serious side effects and dietary restrictions)

Keep in mind that individual response to medications can vary greatly, and this is only an initial assessment. It's crucial to work closely with a healthcare professional when determining the most appropriate treatment for a patient.

Vignette #3

#1

I am not a medical doctor, but I can provide some general information on these medications based on their classes, mechanisms of action, and known side effects. It is important to remember that individual cases can vary, and a qualified healthcare professional should always make the final decision on medication.

1. Best choice for this patient:

- Sertraline: An SSRI with a good efficacy and side effect profile for depression.
- Escitalopram: Another SSRI that could be effective for depression and generally well-tolerated.
- Citalopram: A third SSRI option that could be helpful for this patient's depression.

2. Good choice for this patient:

- Fluoxetine: An SSRI that can be effective for depression but may take longer to see results.
- Vortioxetine: A newer antidepressant with multimodal action, could be helpful if SSRIs aren't effective.
- Duloxetine: An SNRI that may be helpful for depression, but monitor for potential interaction with amoxicillin.
- Mirtazapine: Atypical antidepressant that can be helpful, especially if the patient has insomnia or appetite issues.

3. Less good choice for this patient:

- Bupropion: Could be useful for depression, but not ideal for patients with atrial fibrillation due to potential increased risk of cardiovascular events.
- Venlafaxine: An SNRI that can be effective but may have more side effects and drug interactions.
- Fluvoxamine: An SSRI that is generally less preferred for depression, but may be an option if others aren't effective.

4. Bad choice for this patient, or contraindicated:

- Tricyclic antidepressants (Amitriptyline, Clomipramine, Desipramine, Imipramine, Nortriptyline, Protriptyline): Not ideal due to higher side effect profile and potential to worsen atrial fibrillation.
- Paroxetine: Already tried and reported as ineffective for this patient.
- MAOIs (Isocarboxazid, Phenelzine, Tranylcypromine): Generally reserved for treatment-resistant cases due to dietary restrictions, potential drug interactions, and side effects.
- Milnacipran: Primarily used for fibromyalgia, not typically prescribed for depression.

Please consult with a healthcare professional to determine the best course of action for this patient, as individual factors may influence the decision-making process.

#2

Best choice for this patient:

1. Escitalopram
2. Sertraline
3. Fluoxetine

Good choice for this patient: 4. Citalopram

5. Duloxetine
6. Venlafaxine
7. Vortioxetine

Less good choice for this patient: 8. Bupropion (due to the risk of exacerbating atrial fibrillation)

9. Mirtazapine (may increase appetite and weight, which may not be ideal for someone with atrial fibrillation)
10. Nortriptyline (due to anticholinergic side effects and potential cardiac effects)
11. Desipramine (due to anticholinergic side effects and potential cardiac effects)
12. Fluvoxamine (not as commonly used for depression as other SSRIs)
13. Milnacipran (more commonly used for fibromyalgia, not as well-established for depression)

Bad choice for this patient, or contraindicated: 14. Amitriptyline (due to anticholinergic side effects and potential cardiac effects)

15. Clomipramine (due to anticholinergic side effects and potential cardiac effects)
16. Imipramine (due to anticholinergic side effects and potential cardiac effects)
17. Paroxetine (already tried and not effective)
18. Protriptyline (due to anticholinergic side effects and potential cardiac effects)
19. Isocarboxazid (MAOI with significant dietary restrictions and drug interactions)
20. Phenelzine (MAOI with significant dietary restrictions and drug interactions)
21. Tranylcypromine (MAOI with significant dietary restrictions and drug interactions)

#3

Best choice for this patient:

1. Escitalopram
2. Sertraline
3. Fluoxetine
4. Vortioxetine
5. Duloxetine
6. Venlafaxine
7. Citalopram

Good choice for this patient:

1. Bupropion (Note: Consider potential risk due to atrial fibrillation)
2. Mirtazapine
3. Milnacipran

Less good choice for this patient:

1. Fluvoxamine
2. Nortriptyline
3. Desipramine
4. Imipramine
5. Protriptyline
6. Clomipramine

Bad choice for this patient, or contraindicated:

1. Amitriptyline (due to its anticholinergic effects, which might worsen atopic dermatitis)
2. Paroxetine (already tried without success)
3. Isocarboxazid (due to potential drug interactions with fluconazole)
4. Phenelzine (due to potential drug interactions with fluconazole)
5. Tranylcypromine (due to potential drug interactions with fluconazole)

#4

Best choice for this patient:

- Escitalopram
- Sertraline
- Vortioxetine
- Duloxetine

Good choice for this patient:

- Citalopram
- Fluoxetine
- Fluvoxamine
- Venlafaxine
- Mirtazapine
- Bupropion (though it should be used with caution due to the patient's history of atrial fibrillation)

Less good choice for this patient:

- Nortriptyline
- Desipramine
- Clomipramine
- Imipramine
- Protriptyline
- Milnacipran

Bad choice for this patient, or contraindicated:

- Amitriptyline (due to the patient's history of atrial fibrillation)
- Isocarboxazid (due to potential interactions with clarithromycin)
- Phenelzine (due to potential interactions with clarithromycin)
- Tranylcypromine (due to potential interactions with clarithromycin)
- Paroxetine (since it did not cause a reduction in depression symptoms)

[additional interaction]

*can you explain the interactions with clarithromycin?*

I'm not a doctor, but I can provide you with some general information on the potential interactions between clarithromycin and certain medications.

Clarithromycin is an antibiotic that belongs to the class of macrolides. It is used to treat a variety of bacterial infections. It can interact with several medications due to its effect on the liver enzyme CYP3A4. This enzyme is responsible for the metabolism of a wide range of drugs. When clarithromycin is taken, it can inhibit the activity of CYP3A4, leading to increased levels of certain medications in the bloodstream, potentially causing adverse effects or toxicity.

In the case of monoamine oxidase inhibitors (MAOIs) like isocarboxazid, phenelzine, and tranylcypromine, combining them with clarithromycin can increase the risk of serious side effects, including serotonin syndrome. Serotonin syndrome is a potentially life-threatening condition caused by an excessive accumulation of serotonin in the body. Symptoms can include agitation, confusion, rapid heart rate, high blood pressure, muscle rigidity, and in severe cases, coma or even death.

#5

1. Best choice for this patient:

- Sertraline: An SSRI with fewer drug interactions and generally well-tolerated.
- Escitalopram: Another SSRI with a good safety profile and fewer drug interactions.
- Vortioxetine: An atypical antidepressant with low potential for drug interactions.

2. Good choice for this patient:

- Citalopram: An SSRI, although it has a higher risk of QT prolongation compared to other SSRIs.
- Fluoxetine: An SSRI with a long half-life, which may be helpful, but it has more drug interactions.
- Duloxetine: An SNRI, but may exacerbate atopic dermatitis in some cases.
- Mirtazapine: An atypical antidepressant with low potential for drug interactions, but may cause weight gain.

3. Less good choice for this patient:

- Bupropion: An NDRI, can be helpful in depression but contraindicated in patients with a history of seizures or eating disorders.
- Venlafaxine: An SNRI, with a risk of exacerbating hypertension and potential drug interactions.
- Fluvoxamine: An SSRI primarily used for OCD, but may be useful in depression.

4. Bad choice for this patient, or contraindicated:

- Amitriptyline, Clomipramine, Desipramine, Imipramine, Nortriptyline, Protriptyline: TCAs, which have a higher risk of adverse effects and drug interactions, especially with quinidine.
- Isocarboxazid, Phenelzine, Tranylcypromine: MAOIs, which have a high risk of drug interactions and dietary restrictions.
- Paroxetine: Already tried and did not cause a reduction in depression symptoms.
- Milnacipran: An SNRI primarily used for fibromyalgia, and may not be the most appropriate choice for depression.

Vignette #4

#1

I'm not a doctor, but I can provide you with some general information based on the case you presented. It's important to consult a medical professional for personalized advice.

1. Best choice for this patient:

- Bupropion: May help with depression and has a potential side effect of weight loss, which could be beneficial given her history of obesity.
- Duloxetine: Effective for depression and may also help with diabetic neuropathy, making it a suitable option for Sarah.
- Escitalopram: An SSRI that could be effective for depression and generally has a better side effect profile compared to Paroxetine.
- Sertraline: Another SSRI that might be effective for depression and generally has a better side effect profile compared to Paroxetine.

2. Good choice for this patient:

- Citalopram: An SSRI that may be effective for depression, but has a similar profile to Paroxetine.
- Fluoxetine: An SSRI that may be effective for depression, but has a similar profile to Paroxetine.
- Mirtazapine: Can be effective for depression and may help with sleep, but has a risk of weight gain.
- Vortioxetine: May be effective for depression, but has a similar profile to SSRIs.

3. Less good choice for this patient:

- Amitriptyline: A tricyclic antidepressant that can be effective for depression, but has more side effects and potential for weight gain.
- Clomipramine: A tricyclic antidepressant that can be effective for depression, but has more side effects and potential for weight gain.
- Desipramine: A tricyclic antidepressant that can be effective for depression, but has more side effects and potential for weight gain.
- Fluvoxamine: An SSRI that may be effective for depression, but has a similar profile to Paroxetine.
- Imipramine: A tricyclic antidepressant that can be effective for depression, but has more side effects and potential for weight gain.
- Milnacipran: A serotonin-norepinephrine reuptake inhibitor (SNRI) that can be effective for depression, but has a similar profile to other SNRIs.
- Nortriptyline: A tricyclic antidepressant that can be effective for depression, but has more side effects and potential for weight gain.
- Venlafaxine: An SNRI that can be effective for depression, but has a similar profile to other SNRIs.

4. Bad choice for this patient, or contraindicated:

- Isocarboxazid: A monoamine oxidase inhibitor (MAOI) that can be effective for depression, but has many dietary restrictions and drug interactions, making it less suitable for Sarah.
- Paroxetine: As mentioned, prior treatment with Paroxetine did not cause a reduction in depression symptoms for Sarah.
- Phenelzine: An MAOI that can be effective for depression, but has many dietary restrictions and drug interactions, making it less suitable for Sarah.
- Protriptyline: A tricyclic antidepressant that can be effective for depression, but has more side effects and potential for weight gain.
- Tranylcypromine: An MAOI that can be effective for depression, but has many dietary restrictions and drug interactions, making it less suitable for Sarah.

Remember, it's essential to consult a healthcare professional before making any decisions about medication.

#2

Best choice for this patient:

1. Bupropion: Can help with fatigue, weight management, and is less likely to cause sexual side effects.
2. Duloxetine: Can help with both depression and diabetic neuropathy pain.
3. Vortioxetine: May be effective in treating depression with a lower risk of weight gain and sexual side effects.

Good choice for this patient:

1. Citalopram: An SSRI that could be considered if other first-line options are not effective.
2. Fluoxetine: Another SSRI that might be beneficial.
3. Sertraline: An SSRI that may be tried if other options are not successful.
4. Venlafaxine: A serotonin-norepinephrine reuptake inhibitor (SNRI) that could be helpful in managing depression.
5. Mirtazapine: May be useful for treating depression and can help with sleep, but may cause weight gain.

Less good choice for this patient:

1. Escitalopram: Already tried and not effective for this patient.
2. Paroxetine: An SSRI with a higher risk of weight gain and sexual side effects compared to other SSRIs.
3. Fluvoxamine: An SSRI typically used for obsessive-compulsive disorder (OCD) rather than depression.
4. Nortriptyline: A tricyclic antidepressant (TCA) with a higher risk of side effects than SSRIs and SNRIs.
5. Desipramine: Another TCA with a higher risk of side effects compared to SSRIs and SNRIs.
6. Milnacipran: An SNRI typically used for fibromyalgia, not specifically for depression.

Bad choice for this patient, or contraindicated:

1. Amitriptyline: A TCA with a higher risk of weight gain and adverse side effects.
2. Clomipramine: A TCA typically used for OCD, with a higher risk of side effects.
3. Imipramine: Another TCA with a higher risk of side effects.
4. Protriptyline: A TCA with a higher risk of side effects compared to SSRIs and SNRIs.
5. Isocarboxazid: A monoamine oxidase inhibitor (MAOI) with significant dietary restrictions and drug interactions.
6. Phenelzine: Another MAOI with significant dietary restrictions and drug interactions.
7. Tranylcypromine: Yet another MAOI with significant dietary restrictions and drug interactions.

#3

Best choice for this patient:

1. Bupropion: Often used to treat depression and may help with fatigue; has less risk of weight gain compared to some other antidepressants.
2. Duloxetine: An SNRI that can be helpful in treating depression and may also help with diabetes-related neuropathy.
3. Escitalopram: An SSRI with a relatively lower risk of weight gain compared to other SSRIs.

Good choice for this patient: 4. Sertraline: An SSRI that is commonly used to treat depression.

5. Citalopram: Another SSRI, but may have a slightly higher risk of weight gain compared to escitalopram or sertraline.
6. Venlafaxine: An SNRI, similar to duloxetine, but with potentially more side effects.
7. Vortioxetine: A newer atypical antidepressant that might be effective in treating depression.

Less good choice for this patient:

8. Mirtazapine: An atypical antidepressant that is effective in treating depression, but has a high risk of weight gain.

9. Paroxetine: An SSRI with a higher risk of weight gain compared to other SSRIs.
10. Nortriptyline: A TCA with less side effects than amitriptyline, but still not a first-line treatment.

Bad choice for this patient, or contraindicated: 11. Amitriptyline: A TCA with a higher risk of weight gain and other side effects, not typically used as a first-line treatment for depression.

12. Clomipramine: A TCA with more side effects, typically used for obsessive-compulsive disorder.
13. Desipramine: A TCA, not usually used as a first-line treatment for depression due to side effects.
14. Fluoxetine: Already tried without success in this patient.
15. Fluvoxamine: An SSRI primarily used for obsessive-compulsive disorder.
16. Imipramine: A TCA, not usually used as a first-line treatment for depression due to side effects.
17. Isocarboxazid: An MAOI, typically reserved for treatment-resistant depression due to dietary restrictions and potential interactions.
18. Milnacipran: An SNRI approved for fibromyalgia, not a first-line treatment for depression.
19. Phenelzine: An MAOI, typically reserved for treatment-resistant depression due to dietary restrictions and potential interactions.
20. Protriptyline: A TCA with side effects, not usually used as a first-line treatment for depression.
21. Tranylcypromine: An MAOI, typically reserved for treatment-resistant depression due to dietary restrictions and potential interactions.

#4

1. Best choice for this patient:

- Bupropion: This medication may be particularly beneficial for Steve because it can help with both depression and fatigue, and it has a lower risk of weight gain compared to some other antidepressants.
- Duloxetine: This medication could be helpful for Steve as it is an SNRI and has shown efficacy in treating depression, and it may have some benefits for diabetic peripheral neuropathy.

2. Good choice for this patient:

- Citalopram
- Escitalopram
- Sertraline
- Venlafaxine
- Vortioxetine

These medications are all commonly used to treat depression and may have a better side effect profile compared to other options. While fluoxetine was not effective for Steve, other SSRIs or SNRIs might still be helpful.

3. Less good choice for this patient:

- Amitriptyline
- Clomipramine
- Desipramine
- Imipramine
- Nortriptyline
- Paroxetine
- Protriptyline

These medications are tricyclic antidepressants (TCAs) or have more side effects compared to other options. They might be considered if other options have been exhausted, but they may not be the first line of treatment for this patient.

4. Bad choice for this patient, or contraindicated:

- Fluoxetine: As mentioned, fluoxetine has already been tried without success in this case, so it's not a suitable option.
- Fluvoxamine: While it's an SSRI, it has a higher risk of drug-drug interactions and might not be the best choice for this patient.
- Isocarboxazid
- Phenelzine
- Tranylcypromine

These medications are monoamine oxidase inhibitors (MAOIs) and generally have more side effects and dietary restrictions compared to other antidepressants. They are typically reserved for treatment-resistant cases and may not be appropriate for this patient at this time.

5. Insufficient information:

- Milnacipran: This medication is primarily indicated for fibromyalgia and has less evidence for treating depression.
- Mirtazapine: While it can be effective for depression, it has a higher risk of weight gain, which may not be ideal for a patient with obesity.

Please remember that the information provided is general and should not be used as medical advice. A qualified healthcare professional should always be consulted for individualized advice.

#5

1. Best choice for this patient:
   - Bupropion: Can help with depression and may also aid in weight loss.
   - Duloxetine: Effective for depression and may help with diabetes-related neuropathic pain.
   - Vortioxetine: An effective antidepressant with a low risk of weight gain.
   - Escitalopram: Effective for depression with fewer side effects than Paroxetine.
2. Good choice for this patient:
   - Sertraline: Effective for depression with a lower risk of weight gain than some other SSRIs.
   - Citalopram: Similar to Escitalopram and Sertraline, but may have slightly more side effects.
   - Venlafaxine: An SNRI that can be effective for depression, but may cause weight gain in some patients.
   - Mirtazapine: Effective for depression and may improve sleep, but may cause weight gain.
   - Fluoxetine: An SSRI that can be effective for depression, but may not be as effective as other SSRIs in this case.
3. Less good choice for this patient:
   - Nortriptyline: A TCA with fewer side effects than Amitriptyline, but still carries risks.
   - Desipramine: Similar to Nortriptyline, but with a slightly different side effect profile.
   - Milnacipran: An SNRI with fewer data on its efficacy in depression, and potential for weight gain.
   - Fluvoxamine: An SSRI primarily used for OCD, but may be less effective for depression.
4. Bad choice for this patient, or contraindicated:
   - Paroxetine: Already tried without success.
   - Amitriptyline: A TCA with higher risk of weight gain and other side effects.
   - Clomipramine: A TCA primarily used for OCD, with a higher risk of side effects.
   - Imipramine: A TCA with a higher risk of weight gain and side effects.
   - Protriptyline: A TCA with a higher risk of side effects and weight gain.
   - Isocarboxazid: An MAOI with significant dietary restrictions and potential side effects.
   - Phenelzine: An MAOI with significant dietary restrictions and potential side effects.
   - Tranylcypromine: An MAOI with significant dietary restrictions and potential side effects.

Please note that this is only an analysis based on the information provided and should not be considered medical advice. It is important to consult a healthcare professional for a comprehensive evaluation and personalized treatment plan.

Vignette #5

#1

It's important to note that I am an AI language model and not a licensed psychiatrist, so any suggestions I provide should be taken as informational only and should not replace professional medical advice. However, based on the information provided, here's a rough rating of the medications for the case presented:

Best choice for this patient:

1. Duloxetine
2. Venlafaxine
3. Vortioxetine
4. Escitalopram
5. Sertraline
6. Fluoxetine
7. Mirtazapine
8. Paroxetine

Good choice for this patient:

9. Milnacipran

10. Fluvoxamine

Less good choice for this patient:

11. Bupropion (potential risk of seizures, given the patient's history of seizure disorder)

12. Nortriptyline
13. Desipramine
14. Protriptyline
15. Imipramine
16. Clomipramine
17. Amitriptyline (higher risk of side effects)

Bad choice for this patient, or contraindicated: 18. Citalopram (already tried, did not help with depression symptoms)

19. Isocarboxazid (MAOI, higher risk of drug interactions and side effects)
20. Phenelzine (MAOI, higher risk of drug interactions and side effects)
21. Tranylcypromine (MAOI, higher risk of drug interactions and side effects)

Please remember that this is a rough rating and should not replace the judgment of a trained professional. The specific details of the case and the patient's individual needs should be considered when making a final decision about medication.

#2

Best choice for this patient:

1. Sertraline
2. Escitalopram
3. Citalopram
4. Fluoxetine
5. Vortioxetine

Good choice for this patient: 6. Venlafaxine

7. Duloxetine
8. Milnacipran
9. Mirtazapine

Less good choice for this patient: 10. Desipramine

11. Nortriptyline
12. Protriptyline

Bad choice for this patient, or contraindicated: 13. Bupropion (due to the history of seizure disorder)

14. Clomipramine (risk of lowering seizure threshold)
15. Imipramine (risk of lowering seizure threshold)
16. Amitriptyline (risk of lowering seizure threshold)
17. Paroxetine (already tried without success)
18. Fluvoxamine (may have potential drug interaction with Celecoxib)
19. Isocarboxazid (risk of hypertensive crisis with certain drug interactions)
20. Phenelzine (risk of hypertensive crisis with certain drug interactions)
21. Tranylcypromine (risk of hypertensive crisis with certain drug interactions)

#3

Best choice for this patient:

1. Sertraline
2. Escitalopram
3. Fluoxetine
4. Paroxetine
5. Venlafaxine
6. Duloxetine
7. Vortioxetine
8. Mirtazapine

Good choice for this patient:

1. Bupropion (note that there may be a potential risk of seizures in a patient with a seizure disorder, so use cautiously)
2. Milnacipran

Less good choice for this patient:

1. Desipramine
2. Nortriptyline
3. Protriptyline
4. Clomipramine
5. Imipramine
6. Fluvoxamine

Bad choice for this patient, or contraindicated:

1. Amitriptyline (due to higher risk of seizures)
2. Citalopram (already tried without success)
3. Isocarboxazid (due to risk of hypertensive crisis and potential interactions with other medications)
4. Phenelzine (due to risk of hypertensive crisis and potential interactions with other medications)
5. Tranylcypromine (due to risk of hypertensive crisis and potential interactions with other medications)

#4

Best choice for this patient:

1. Sertraline
2. Fluoxetine
3. Vortioxetine
4. Duloxetine
5. Venlafaxine

Good choice for this patient: 6. Citalopram

7. Paroxetine
8. Mirtazapine
9. Milnacipran

Less good choice for this patient: 10. Bupropion (potential risk with seizure disorder)

11. Nortriptyline
12. Desipramine
13. Clomipramine
14. Imipramine
15. Protriptyline
16. Escitalopram (since it did not cause a reduction in depression symptoms)

Bad choice for this patient, or contraindicated: 17. Amitriptyline (potential risk with seizure disorder)

18. Fluvoxamine (less evidence for its effectiveness in treating major depressive disorder)
19. Isocarboxazid (MAOI with many interactions, used only if other options are ineffective)
20. Phenelzine (MAOI with many interactions, used only if other options are ineffective)
21. Tranylcypromine (MAOI with many interactions, used only if other options are ineffective)

#5

Best choice for this patient:

1. Escitalopram
2. Sertraline
3. Citalopram
4. Vortioxetine
5. Duloxetine
6. Venlafaxine

Good choice for this patient:

1. Mirtazapine
2. Paroxetine
3. Fluvoxamine
4. Milnacipran

Less good choice for this patient:

1. Bupropion (due to seizure disorder history)
2. Desipramine
3. Nortriptyline
4. Protriptyline

Bad choice for this patient, or contraindicated:

1. Amitriptyline (due to seizure disorder history)
2. Clomipramine (due to seizure disorder history)
3. Imipramine (due to seizure disorder history)
4. Isocarboxazid (due to potential interaction with seizure medications)
5. Phenelzine (due to potential interaction with seizure medications)
6. Tranylcypromine (due to potential interaction with seizure medications)
7. Fluoxetine (as previously tried without success)

[alternate prompt for Vignette #1]

Vignette #1, alternate prompt:

You are the senior psychiatrist who is teaching psychiatry residents. When they describe a case to them, you tell them which medicines you think would be most helpful for the case. These are not real people. Please rank the 5 best medicines for each case and explain why you choose each one. The first case they present is: {case_text}

I am not a doctor, but based on the information provided, here is a hypothetical list of medications that could be considered for Sarah's case. It is important to consult with a healthcare professional before making any decisions about medications.

1. Sertraline (Zoloft) - Sertraline is an SSRI that could be considered for treating depression. It may be chosen as the first option because it has a lower likelihood of causing arrhythmia than other SSRIs like Paroxetine. It is also considered safe in patients with hypertension and diabetes, but careful monitoring would be necessary due to her existing conditions.
2. Bupropion (Wellbutrin) - Bupropion is an atypical antidepressant that can be used to treat depression. It has a lower risk of causing cardiovascular side effects compared to other antidepressants, which may be beneficial for someone with a history of arrhythmia and hypertensive heart disease. Additionally, bupropion has been shown to have positive effects on glycemic control in patients with diabetes.
3. Venlafaxine (Effexor) - Venlafaxine is an SNRI that could be considered for treating depression. It has been shown to be effective in patients with treatment-resistant depression, and it may have a lower risk of cardiac side effects compared to other antidepressants. However, it is essential to monitor her blood pressure, as it can cause an increase in some patients.
4. Mirtazapine (Remeron) - Mirtazapine is an atypical antidepressant that may be used in cases where other antidepressants are not effective or tolerated. It has a low risk of causing arrhythmia, but it may lead to weight gain and an increased appetite, which could be problematic for a patient with diabetes.
5. Duloxetine (Cymbalta) - Duloxetine is an SNRI that can be considered for treating depression, especially when accompanied by chronic pain. It may have a lower risk of cardiac side effects than other antidepressants. However, it's important to monitor her blood pressure and blood sugar levels, as Duloxetine could potentially exacerbate her existing conditions.

In all cases, it is important to closely monitor Sarah's response to these medications and adjust the treatment plan accordingly. Close collaboration with her primary care physician and a cardiologist would also be essential in managing her comorbid conditions while treating her depression.

Second round of vignettes

Vignette #1

#1:

I'm not a medical doctor, but I can try to help you think through these options based on the information given. Remember to consult a healthcare professional for medical advice. Here's a potential rating of the medications for Sarah's case:

1. "Best choice for this patient":

- Sertraline: It is an SSRI that might be effective for depression and has fewer cardiac side effects.
- Escitalopram: Similar to sertraline, it is an SSRI with a good safety profile and may be effective for depression.
- Duloxetine: A serotonin-norepinephrine reuptake inhibitor (SNRI) that could address both depression and diabetic neuropathy.

2. "Good choice for this patient":

- Citalopram: An SSRI with potential benefits for depression, but less favorable than escitalopram or sertraline due to its potential for QT interval prolongation.
- Fluoxetine: Another SSRI that may be useful for depression, but it has a longer half-life, which can lead to drug interactions.
- Vortioxetine: An atypical antidepressant that could be effective for depression but may have less clinical evidence than other options.

3. "Less good choice for this patient":

- Bupropion: An atypical antidepressant that can be useful for depression, but it could potentially exacerbate arrhythmias.
- Mirtazapine: An atypical antidepressant that might be helpful for depression but can cause weight gain, which may not be ideal given her diabetes.
- Venlafaxine: An SNRI that may be useful for depression but has a higher risk of causing hypertension compared to other options.

4. "Bad choice for this patient, or contraindicated":

- Amitriptyline, Clomipramine, Desipramine, Imipramine, Nortriptyline, Protriptyline: These are tricyclic antidepressants (TCAs) which are generally not recommended due to their potential for cardiac side effects, particularly in patients with arrhythmia and hypertensive heart disease.
- Isocarboxazid, Phenelzine, Tranylcypromine: These are monoamine oxidase inhibitors (MAOIs) which have significant dietary restrictions and drug interactions, making them less suitable for this patient.
- Fluvoxamine: An SSRI that is primarily used for obsessive-compulsive disorder (OCD) and may not be the best choice for depression in this case.
- Milnacipran: An SNRI that is mainly used for fibromyalgia and has less evidence for depression treatment.
- Paroxetine: As mentioned, prior treatment with paroxetine did not cause a reduction in depression symptoms for this patient, making it an unsuitable choice.

#2

Best choice for this patient:

1. Bupropion
2. Sertraline
3. Escitalopram
4. Citalopram
5. Fluoxetine
6. Paroxetine
7. Vortioxetine
8. Duloxetine

Good choice for this patient: 9. Mirtazapine

10. Milnacipran

Less good choice for this patient: 11. Desipramine

12. Nortriptyline
13. Protriptyline
14. Fluvoxamine
15. Venlafaxine (already tried without success)

Bad choice for this patient, or contraindicated: 16. Amitriptyline (due to arrhythmia and hypertensive heart disease)

17. Clomipramine (due to arrhythmia and hypertensive heart disease)
18. Imipramine (due to arrhythmia and hypertensive heart disease)
19. Isocarboxazid (due to hypertensive heart disease)
20. Phenelzine (due to hypertensive heart disease)
21. Tranylcypromine (due to hypertensive heart disease)

#3

1. Best choice for this patient:

- Sertraline: An SSRI with fewer cardiovascular side effects, it may be a better option than citalopram for this patient with a history of arrhythmia and hypertensive heart disease.
- Duloxetine: An SNRI that can help with depression and has been shown to have benefits for diabetic neuropathy, which may be relevant for Sarah.

2. Good choice for this patient:

- Vortioxetine: An atypical antidepressant that has demonstrated efficacy in treating depression and a lower risk of cardiovascular side effects.
- Venlafaxine: An SNRI with demonstrated efficacy in treating depression, but it should be used cautiously in patients with cardiovascular issues.
- Escitalopram: Another SSRI similar to citalopram, but potentially with fewer cardiovascular side effects. However, since citalopram did not help the patient, escitalopram may not be the best option.
- Mirtazapine: An atypical antidepressant that may be helpful for depression and sleep, but should be used cautiously in patients with diabetes due to potential weight gain.

3. Less good choice for this patient:

- Bupropion: It can be helpful for depression, but it may not be the best choice for someone with a history of arrhythmia or hypertensive heart disease, as it may raise blood pressure.
- Fluoxetine, Paroxetine, and Fluvoxamine: SSRIs similar to citalopram, but since citalopram did not help the patient, these may not be the best options.

4. Bad choice for this patient, or contraindicated:

- Amitriptyline, Clomipramine, Desipramine, Imipramine, Nortriptyline, and Protriptyline: These tricyclic antidepressants are generally not recommended for patients with a history of arrhythmia or heart disease due to their potential cardiovascular side effects.
- Isocarboxazid, Phenelzine, and Tranylcypromine: These MAOIs have significant dietary and drug interactions, including hypertensive crises, and are generally not first-line treatments for depression. They are not ideal for patients with heart disease.
- Milnacipran: An SNRI with limited data on its efficacy for depression, and it may not be the best choice for someone with cardiovascular issues.
- Citalopram: As mentioned in the case presentation, citalopram did not cause a reduction in depression symptoms for this patient. Additionally, it may not be the best choice for someone with a history of arrhythmia.

#4

1. Best choice for this patient:

- Sertraline: An SSRI that may be more tolerable for patients with heart disease and diabetes.

2. Good choice for this patient:

- Citalopram: An SSRI with a lower risk of drug-drug interactions compared to other SSRIs.
- Escitalopram: An SSRI that is typically well-tolerated and has a lower risk of drug-drug interactions.
- Fluoxetine: An SSRI with a long half-life, which may be helpful in patients with compliance issues.
- Vortioxetine: A multimodal antidepressant that may offer additional benefits in cognitive functioning.

3. Less good choice for this patient:

- Bupropion: An atypical antidepressant, but it may increase the risk of arrhythmia.
- Duloxetine: An SNRI that may be useful for depression but might worsen blood pressure.
- Fluvoxamine: An SSRI with higher risk of drug-drug interactions, especially with ritonavir.
- Mirtazapine: An atypical antidepressant with potential for weight gain and sedation, which may not be ideal for a diabetic patient.
- Venlafaxine: An SNRI with potential to exacerbate blood pressure issues.

4. Bad choice for this patient, or contraindicated:

- Amitriptyline: A tricyclic antidepressant (TCA) with a higher risk of arrhythmia and worsening blood pressure.
- Clomipramine: A TCA with a higher risk of arrhythmia and worsening blood pressure.
- Desipramine: A TCA with a higher risk of arrhythmia and worsening blood pressure.
- Imipramine: A TCA with a higher risk of arrhythmia and worsening blood pressure.
- Isocarboxazid: An MAOI with significant drug-drug interactions and dietary restrictions.
- Milnacipran: An SNRI not commonly used for depression and with potential to exacerbate blood pressure issues.
- Nortriptyline: A TCA with a higher risk of arrhythmia and worsening blood pressure.
- Paroxetine: Previously tried and failed to reduce depression symptoms.
- Phenelzine: An MAOI with significant drug-drug interactions and dietary restrictions.
- Protriptyline: A TCA with a higher risk of arrhythmia and worsening blood pressure.
- Tranylcypromine: An MAOI with significant drug-drug interactions and dietary restrictions.

#5

1. Best choice for this patient:

- Sertraline: This SSRI has a low risk of cardiotoxicity and may be suitable for someone with cardiac conditions.
- Escitalopram: This SSRI is also considered safe for patients with cardiac issues and has shown efficacy in treating depression.

2. Good choice for this patient:

- Citalopram: This SSRI has a lower risk of cardiotoxicity but might require monitoring due to potential QT interval prolongation.
- Fluoxetine: Another SSRI that might be suitable for this patient, but potential interactions with fluconazole should be considered.
- Vortioxetine: This atypical antidepressant may be an option with lower risk for cardiac issues.

3. Less good choice for this patient:

- Bupropion: While it's an atypical antidepressant with low cardiotoxicity risk, it can increase blood pressure, which may not be ideal for hypertensive patients.
- Duloxetine: This SNRI may be helpful for depression but has the potential to affect blood pressure.
- Mirtazapine: This atypical antidepressant has lower cardiotoxicity risk but may cause weight gain, which isn't ideal for diabetic patients.
- Paroxetine: This SSRI has a lower risk of cardiotoxicity, but it might not be the first choice due to side effects and potential drug interactions.

4. Bad choice for this patient, or contraindicated:

- Amitriptyline, Clomipramine, Desipramine, Imipramine, Nortriptyline, and Protriptyline: These tricyclic antidepressants have higher cardiotoxicity risk and should be avoided in patients with cardiac issues.
- Isocarboxazid, Phenelzine, and Tranylcypromine: These MAOIs have potential for significant drug interactions and hypertensive crises, making them less suitable for this patient.
- Milnacipran: This SNRI has potential to affect blood pressure and heart rate and might not be the best choice for someone with cardiac conditions.
- Venlafaxine: As it was previously tried and didn't show effectiveness, it may not be suitable for this patient.

Vignette #2

#1

Best choice for this patient:

1. Clomipramine: It is an effective treatment for both depression and OCD, which Sarah has a history of.
2. Fluvoxamine: Another SSRI that might be effective for depression and OCD.
3. Sertraline: An SSRI that has shown efficacy in treating depression and OCD.

Good choice for this patient: 4. Escitalopram: An SSRI that may be helpful for depression, but not specifically targeted for OCD.

5. Fluoxetine: An SSRI that can be useful in treating depression and has some evidence for OCD treatment.
6. Paroxetine: Another SSRI that can be useful for depression and has some evidence for OCD treatment.
7. Venlafaxine: An SNRI that may be helpful for depression, but not specifically targeted for OCD.
8. Duloxetine: An SNRI that may be helpful for depression, but not specifically targeted for OCD.
9. Vortioxetine: A newer antidepressant that may be helpful for depression, but not specifically targeted for OCD.

Less good choice for this patient: 10. Bupropion: An atypical antidepressant, may be less effective for OCD.

11. Mirtazapine: An atypical antidepressant, may be less effective for OCD.
12. Nortriptyline: A TCA, may be less effective for OCD.
13. Desipramine: A TCA, may be less effective for OCD.

Bad choice for this patient, or contraindicated: 14. Citalopram: Already tried with no reduction in depression symptoms.

15. Amitriptyline: A TCA, may have more side effects and be less effective for OCD.
16. Imipramine: A TCA, may have more side effects and be less effective for OCD.
17. Protriptyline: A TCA, may have more side effects and be less effective for OCD.
18. Isocarboxazid: An MAOI, generally reserved for treatment-resistant cases due to side effects and dietary restrictions.
19. Phenelzine: An MAOI, generally reserved for treatment-resistant cases due to side effects and dietary restrictions.
20. Tranylcypromine: An MAOI, generally reserved for treatment-resistant cases due to side effects and dietary restrictions.
21. Milnacipran: An SNRI primarily used for fibromyalgia, not specifically targeted for depression or OCD.

#2

1. Best choice for this patient:

- Clomipramine: It is an effective medication for both depression and OCD symptoms.
- Sertraline: An SSRI with demonstrated efficacy in both depression and OCD.
- Fluvoxamine: Another SSRI with efficacy in treating both depression and OCD.

2. Good choice for this patient:

- Escitalopram: An SSRI that can be effective for depression and may help with OCD.
- Fluoxetine: An SSRI with potential efficacy in treating depression and OCD.
- Paroxetine: An SSRI with potential efficacy in treating depression and OCD.
- Venlafaxine: An SNRI that can be effective for depression and may help with OCD.
- Duloxetine: An SNRI that can be effective for depression and may help with OCD.
- Vortioxetine: An atypical antidepressant that can be effective for depression, but less evidence for OCD.

3. Less good choice for this patient:

- Bupropion: An atypical antidepressant that can be effective for depression, but not typically used for OCD.
- Mirtazapine: An atypical antidepressant that can be effective for depression, but not typically used for OCD.
- Nortriptyline: A TCA that can be effective for depression, but not typically used for OCD.
- Desipramine: A TCA that can be effective for depression, but not typically used for OCD.

4. Bad choice for this patient, or contraindicated:

- Amitriptyline: A TCA with potential anticholinergic side effects and weight gain, which may not be suitable for this patient.
- Citalopram: The patient has already failed to respond to this medication.
- Imipramine: A TCA with potential anticholinergic side effects and weight gain, which may not be suitable for this patient.
- Isocarboxazid: An MAOI with many potential side effects and dietary restrictions, generally not considered first-line treatment for depression or OCD.
- Milnacipran: An SNRI that is not approved for depression treatment in the US.
- Protriptyline: A TCA with potential anticholinergic side effects and weight gain, which may not be suitable for this patient.
- Phenelzine: An MAOI with many potential side effects and dietary restrictions, generally not considered first-line treatment for depression or OCD.
- Tranylcypromine: An MAOI with many potential side effects and dietary restrictions, generally not considered first-line treatment for depression or OCD.

#3

Best choice for this patient:

1. Clomipramine - It's an effective treatment for OCD and can help with depression as well.
2. Sertraline - Good for both depression and OCD, with a lower side effect profile than clomipramine.
3. Fluvoxamine - Another SSRI that can be effective for both depression and OCD.

Good choice for this patient: 4. Citalopram - An SSRI that can help with depression and potentially OCD.

5. Escitalopram - Another SSRI that can help with depression and potentially OCD.
6. Fluoxetine - An SSRI that can help with depression and potentially OCD.
7. Vortioxetine - An atypical antidepressant that can be effective for depression and may have some benefit for OCD.

Less good choice for this patient: 8. Bupropion - Primarily used for depression but not as effective for OCD.

9. Duloxetine - Effective for depression but not specifically tailored for OCD treatment.
10. Venlafaxine - Effective for depression but not specifically tailored for OCD treatment.
11. Mirtazapine - Effective for depression but not specifically tailored for OCD treatment.
12. Nortriptyline - A tricyclic antidepressant that may be helpful for depression, but not the best option for OCD.
13. Amitriptyline - A tricyclic antidepressant that may be helpful for depression, but not the best option for OCD.
14. Desipramine - A tricyclic antidepressant that may be helpful for depression, but not the best option for OCD.
15. Imipramine - A tricyclic antidepressant that may be helpful for depression, but not the best option for OCD.
16. Protriptyline - A tricyclic antidepressant that may be helpful for depression, but not the best option for OCD.

Bad choice for this patient, or contraindicated: 17. Paroxetine - She has already tried it without success.

18. Isocarboxazid - An MAOI with a high risk of dietary and drug interactions, not the first choice for depression or OCD.
19. Phenelzine - An MAOI with a high risk of dietary and drug interactions, not the first choice for depression or OCD.
20. Tranylcypromine - An MAOI with a high risk of dietary and drug interactions, not the first choice for depression or OCD.
21. Milnacipran - It's an SNRI primarily used for fibromyalgia, not the first choice for depression or OCD.

#4

1. Clomipramine: It's effective for both depression and OCD, which could benefit Sam.

Good choice for this patient: 2. Fluoxetine: An SSRI that can treat both depression and OCD.

3. Sertraline: Another SSRI that can address both depression and OCD.
4. Citalopram: An SSRI that may help with depression, but may be less effective for OCD.
5. Escitalopram: Similar to Citalopram, an SSRI that may help with depression, but may be less effective for OCD.
6. Fluvoxamine: An SSRI that can treat OCD and might also help with depression.
7. Vortioxetine: An atypical antidepressant that may help with depression, but its effectiveness for OCD is not well-established.

Less good choice for this patient: 8. Bupropion: While effective for depression, it's not usually the first choice for OCD.

9. Duloxetine: An SNRI that can treat depression, but its effectiveness for OCD is not well-established.
10. Mirtazapine: An atypical antidepressant that may help with depression, but its effectiveness for OCD is not well-established.
11. Nortriptyline: A tricyclic antidepressant that may help with depression, but its effectiveness for OCD is not well-established.

Bad choice for this patient, or contraindicated: 12. Amitriptyline: A tricyclic antidepressant that may worsen OCD symptoms.

13. Desipramine: A tricyclic antidepressant that may worsen OCD symptoms.
14. Imipramine: A tricyclic antidepressant that may worsen OCD symptoms.
15. Protriptyline: A tricyclic antidepressant that may worsen OCD symptoms.
16. Isocarboxazid: An MAOI, which may have more side effects and interactions, and is generally not a first-line choice.
17. Phenelzine: An MAOI, which may have more side effects and interactions, and is generally not a first-line choice.
18. Tranylcypromine: An MAOI, which may have more side effects and interactions, and is generally not a first-line choice.
19. Milnacipran: An SNRI that is not approved for depression treatment in the United States and may not be effective for OCD.
20. Paroxetine: Known for having more drug-drug interactions and a shorter half-life, which might cause withdrawal symptoms.
21. Venlafaxine: Since it didn't help Sam in the past, it's not a good choice to retry.

Please remember to consult with a healthcare professional for accurate advice on medications.

#5

Best choice for this patient:

1. Clomipramine - Has been shown to be effective for both depression and OCD symptoms.
2. Fluoxetine - SSRI with a good track record for treating depression and OCD.
3. Sertraline - Another SSRI that may be effective in treating both depression and OCD.

Good choice for this patient: 4. Citalopram - SSRI that may help with depression, but with less evidence for OCD.

5. Escitalopram - Similar to citalopram, with potential benefits for depression.
6. Fluvoxamine - SSRI often used for OCD, but may also help with depression.
7. Paroxetine - SSRI with potential benefits for depression and some evidence for OCD.
8. Vortioxetine - Multimodal antidepressant that could help with depression.

Less good choice for this patient: 9. Bupropion - May help with depression but not specifically effective for OCD.

10. Duloxetine - SNRI that could help with depression, but less evidence for OCD.
11. Mirtazapine - Atypical antidepressant that may help with depression, but not specifically for OCD.
12. Nortriptyline - TCA with some potential for treating depression, but not specifically for OCD.

Bad choice for this patient, or contraindicated: 13. Amitriptyline - TCA with more side effects, not specifically effective for OCD.

14. Desipramine - TCA with more side effects, not specifically effective for OCD.
15. Imipramine - TCA with more side effects, not specifically effective for OCD.
16. Isocarboxazid - MAOI with dietary restrictions and potential drug interactions.
17. Milnacipran - SNRI with less evidence for treating depression and not specifically for OCD.
18. Phenelzine - MAOI with dietary restrictions and potential drug interactions.
19. Protriptyline - TCA with more side effects, not specifically effective for OCD.
20. Tranylcypromine - MAOI with dietary restrictions and potential drug interactions.
21. Venlafaxine - As mentioned, prior treatment did not cause a reduction in depression symptoms.

Vignette #3

#1

1. Escitalopram
2. Sertraline
3. Fluoxetine
4. Vortioxetine

Good choice for this patient: 5. Citalopram

6. Duloxetine
7. Venlafaxine
8. Mirtazapine

Less good choice for this patient: 9. Bupropion (due to history of atrial fibrillation)

10. Fluvoxamine
11. Milnacipran
12. Nortriptyline

Bad choice for this patient, or contraindicated: 13. Amitriptyline (due to history of atrial fibrillation and atopic dermatitis)

14. Clomipramine (due to history of atrial fibrillation)
15. Desipramine (due to history of atrial fibrillation)
16. Imipramine (due to history of atrial fibrillation)
17. Isocarboxazid (due to potential interaction with amoxicillin)
18. Paroxetine (ineffective in the past)
19. Phenelzine (due to potential interaction with amoxicillin)
20. Protriptyline (due to history of atrial fibrillation)
21. Tranylcypromine (due to potential interaction with amoxicillin)

#2

1. Best choice for this patient:

- Escitalopram
- Sertraline
- Citalopram
- Fluoxetine
- Vortioxetine

2. Good choice for this patient:

- Bupropion (note: use cautiously due to atrial fibrillation)
- Duloxetine
- Paroxetine
- Mirtazapine
- Milnacipran

3. Less good choice for this patient:

- Fluvoxamine
- Desipramine
- Nortriptyline
- Protriptyline
- Imipramine

4. Bad choice for this patient, or contraindicated:

- Amitriptyline (due to potential for cardiac issues, particularly in patients with atrial fibrillation)
- Clomipramine (due to potential for cardiac issues)
- Isocarboxazid (potential for drug interactions, contraindicated with fluconazole)
- Phenelzine (potential for drug interactions, contraindicated with fluconazole)
- Tranylcypromine (potential for drug interactions, contraindicated with fluconazole)
- Venlafaxine (ineffective in the past)

Please remember that this is only general guidance and that individual patient circumstances may require different considerations. Always consult a medical professional for personalized advice.

#3

1. Best choice for this patient:
   - Escitalopram
   - Sertraline
   - Citalopram
   - Fluoxetine
2. Good choice for this patient:
   - Duloxetine
   - Vortioxetine
   - Paroxetine
   - Mirtazapine
3. Less good choice for this patient:
   - Bupropion (potential concern due to smoking and atrial fibrillation)
   - Milnacipran
   - Fluvoxamine
   - Nortriptyline
   - Desipramine
   - Protriptyline
4. Bad choice for this patient, or contraindicated:
   - Amitriptyline (potential for cardiac side effects with atrial fibrillation)
   - Clomipramine (potential for cardiac side effects with atrial fibrillation)
   - Imipramine (potential for cardiac side effects with atrial fibrillation)
   - Isocarboxazid (potential for interactions with amoxicillin and other side effects)
   - Phenelzine (potential for interactions with amoxicillin and other side effects)
   - Tranylcypromine (potential for interactions with amoxicillin and other side effects)
   - Venlafaxine (ineffective in prior treatment)

#4

1. Escitalopram
2. Sertraline
3. Citalopram
4. Fluoxetine
5. Paroxetine

Good choice for this patient: 6. Vortioxetine

7. Duloxetine
8. Mirtazapine
9. Bupropion

Less good choice for this patient: 10. Milnacipran

11. Fluvoxamine
12. Nortriptyline
13. Desipramine

Bad choice for this patient, or contraindicated: 14. Amitriptyline (due to atrial fibrillation risk)

15. Clomipramine (due to atrial fibrillation risk)
16. Imipramine (due to atrial fibrillation risk)
17. Protriptyline (due to atrial fibrillation risk)
18. Isocarboxazid (due to potential drug interactions)
19. Phenelzine (due to potential drug interactions)
20. Tranylcypromine (due to potential drug interactions)
21. Venlafaxine (already tried without success)

#5

1. Sertraline
2. Escitalopram
3. Citalopram
4. Vortioxetine
5. Duloxetine
6. Venlafaxine
7. Fluoxetine

Good choice for this patient: 8. Mirtazapine

9. Bupropion (be cautious due to smoking history and atrial fibrillation)
10. Fluvoxamine

Less good choice for this patient: 11. Milnacipran

12. Nortriptyline
13. Desipramine
14. Protriptyline

Bad choice for this patient, or contraindicated: 15. Paroxetine (already tried, no reduction in symptoms)

16. Clomipramine (potential drug interaction with ritonavir)
17. Amitriptyline (potential drug interaction with ritonavir)
18. Imipramine (potential drug interaction with ritonavir)
19. Isocarboxazid (potential drug interaction with ritonavir)
20. Phenelzine (potential drug interaction with ritonavir)
21. Tranylcypromine (potential drug interaction with ritonavir)

Vignette #4

#1

Best choice for this patient:

1. Bupropion
2. Duloxetine
3. Escitalopram
4. Fluoxetine
5. Sertraline
6. Venlafaxine
7. Vortioxetine

Good choice for this patient:

1. Citalopram
2. Desipramine
3. Fluvoxamine
4. Mirtazapine
5. Nortriptyline

Less good choice for this patient:

1. Amitriptyline
2. Clomipramine
3. Imipramine
4. Milnacipran
5. Paroxetine (already tried without success)
6. Protriptyline

Bad choice for this patient, or contraindicated:

1. Isocarboxazid
2. Phenelzine
3. Tranylcypromine

#2

1. Best choice for this patient:

- Bupropion: May improve depressive symptoms without causing weight gain, which could be beneficial for a patient with obesity and diabetes.
- Duloxetine: An SNRI that could potentially be more effective than Venlafaxine for this patient, and can also help with diabetic neuropathic pain.
- Escitalopram: An SSRI that might be more effective than Citalopram, with a similar side effect profile.
- Sertraline: An SSRI that could be effective for depressive symptoms and has a lower risk of weight gain.

2. Good choice for this patient:

- Citalopram: An SSRI that may be effective in treating depression, but could cause weight gain.
- Fluoxetine: Another SSRI that may be effective for treating depression but could cause weight gain.
- Vortioxetine: An atypical antidepressant that may be effective in treating depression and has a lower risk of weight gain.

3. Less good choice for this patient:

- Mirtazapine: An atypical antidepressant that may be effective for depression, but is associated with weight gain.
- Paroxetine: An SSRI that may be effective for depression but has a higher risk of weight gain and more pronounced side effects compared to other SSRIs.
- Fluvoxamine: An SSRI with more limited data on its effectiveness for depression compared to other SSRIs, and may cause weight gain.
- Desipramine: A TCA with potential benefits, but could worsen diabetes and obesity.

4. Bad choice for this patient, or contraindicated:

- Amitriptyline: A TCA that may worsen obesity and has potential interactions with diabetes.
- Clomipramine: A TCA with similar concerns to Amitriptyline, and a higher risk of weight gain.
- Imipramine: A TCA with similar concerns to Amitriptyline and Clomipramine.
- Nortriptyline: A TCA with similar concerns to other TCAs, and potential interactions with diabetes.
- Protriptyline: A TCA with similar concerns to other TCAs, and potential interactions with diabetes.
- Isocarboxazid: An MAOI with potential dietary restrictions and drug interactions, making it a less favorable choice.
- Phenelzine: An MAOI with similar concerns as Isocarboxazid.
- Tranylcypromine: An MAOI with similar concerns as other MAOIs.
- Venlafaxine: Previously tried without success.
- Milnacipran: An SNRI primarily indicated for fibromyalgia, with limited data on its use for depression.

#3

Best choice for this patient:

1. Bupropion: As an atypical antidepressant, it may be helpful in treating depression without causing weight gain, and might even promote weight loss. Also, it is known for not worsening fatigue.
2. Duloxetine: As an SNRI, it can be effective for depression and has been studied for use in diabetic peripheral neuropathy, which may be relevant given Sarah's diabetes.

Good choice for this patient:

1. Citalopram: An SSRI that could be considered as an alternative to Escitalopram, which didn't work for Sarah.
2. Sertraline: Another SSRI that could be considered.
3. Venlafaxine: An SNRI that may be effective in treating depression.
4. Vortioxetine: A newer antidepressant with a multimodal mechanism, it may be helpful in cases where other antidepressants haven't been effective.

Less good choice for this patient:

1. Fluoxetine: An SSRI that may cause weight gain or increased fatigue.
2. Paroxetine: An SSRI that can cause weight gain and fatigue, which could worsen Sarah's existing issues.
3. Mirtazapine: Known to cause weight gain and sedation, which might not be ideal for Sarah.
4. Nortriptyline: A TCA that can cause weight gain, which could be an issue in this case.

Bad choice for this patient, or contraindicated:

1. Amitriptyline: A TCA with a higher risk of weight gain and sedation.
2. Clomipramine: A TCA with similar risks as Amitriptyline.
3. Desipramine: A TCA with potential weight gain and sedation risks.
4. Escitalopram: Already tried without success.
5. Fluvoxamine: An SSRI primarily used for OCD, not the best choice for this case.
6. Imipramine: A TCA with similar risks as Amitriptyline.
7. Isocarboxazid: An MAOI with potential dietary restrictions and interactions.
8. Milnacipran: An SNRI primarily used for fibromyalgia, not the best choice for this case.
9. Phenelzine: An MAOI with potential dietary restrictions and interactions.
10. Protriptyline: A TCA with potential weight gain and sedation risks.
11. Tranylcypromine: An MAOI with potential dietary restrictions and interactions.

#4

Best choice for this patient:

1. Bupropion: It is effective in treating depression and could also help with weight loss, which might be beneficial for Samantha considering her obesity.

Good choices for this patient: 2. Duloxetine: Effective for depression and can also help with diabetes-related pain, which could be an additional benefit for Samantha.

3. Escitalopram: It is an SSRI with fewer side effects and can be effective for depression.
4. Fluoxetine: Another SSRI that can be helpful in treating depression.
5. Sertraline: This SSRI is also effective in treating depression and has fewer side effects.

Less good choices for this patient: 6. Citalopram: An SSRI that can be effective for depression, but may have slightly more side effects than others in its class.

7. Fluvoxamine: This SSRI can be helpful for depression, but it's not typically a first-line treatment.
8. Paroxetine: An SSRI that can be effective for depression, but has a higher risk of side effects and weight gain.
9. Mirtazapine: Effective for depression, but may cause weight gain, which could worsen Samantha's obesity.
10. Vortioxetine: Effective for depression, but not as well-studied as other options and may have higher costs.
11. Milnacipran: A SNRI that could be useful for depression, but it's not typically a first-line treatment.

Bad choices for this patient, or contraindicated: 12. Amitriptyline: A tricyclic antidepressant (TCA) with significant side effects and potential for weight gain, which could worsen Samantha's obesity.

13. Clomipramine: A TCA with high side effects and potential for weight gain.
14. Desipramine: A TCA with significant side effects and not typically recommended as first-line treatment for depression.
15. Imipramine: Another TCA with considerable side effects and potential for weight gain.
16. Nortriptyline: A TCA with similar concerns as other TCAs, including side effects and potential weight gain.
17. Protriptyline: A TCA with significant side effects and not typically recommended for depression.
18. Isocarboxazid: An MAOI with potentially serious side effects and dietary restrictions, typically reserved for treatment-resistant cases.
19. Phenelzine: Another MAOI with similar concerns as Isocarboxazid.
20. Tranylcypromine: An MAOI with potentially serious side effects and dietary restrictions, typically reserved for treatment-resistant cases.
21. Venlafaxine: As Samantha has already been treated with Venlafaxine without a reduction in depression symptoms, it would not be a good choice for her.

#5

For Sam, considering his history of diabetes, obesity, and non-response to Paroxetine, these medications could be generally rated as follows:

Best choice for this patient:

1. Bupropion: Its activating properties can help with fatigue and it has a lower risk of weight gain.
2. Duloxetine: It can help with both depression and has some benefits for managing pain related to diabetes.
3. Vortioxetine: It may improve cognitive function and is generally weight neutral.

Good choice for this patient: 4. Escitalopram

5. Fluoxetine
6. Sertraline
7. Citalopram: These SSRIs are generally well-tolerated and effective for depression, though they might not be as helpful for fatigue.
8. Venlafaxine: It is an SNRI and can be effective for depression, but may have some side effects related to weight gain.

Less good choice for this patient: 9. Mirtazapine: Although effective for depression, it may cause weight gain.

10. Nortriptyline
11. Desipramine: These TCA medications can be effective but have more side effects than newer options.
12. Milnacipran: This SNRI has less data supporting its use for depression, but might still be considered.

Bad choice for this patient, or contraindicated: 13. Paroxetine: Already tried without success.

14. Amitriptyline
15. Clomipramine
16. Imipramine
17. Protriptyline: These TCAs have significant side effects and may not be suitable given Sam's health concerns.
18. Fluvoxamine: Mainly used for OCD, less evidence for depression treatment.
19. Isocarboxazid
20. Phenelzine
21. Tranylcypromine: These MAOIs have significant dietary restrictions and interactions, and may not be suitable for someone with diabetes.

Vignette #5

#1

Best choice for this patient:

1. Sertraline
2. Escitalopram
3. Venlafaxine
4. Duloxetine
5. Vortioxetine

Good choice for this patient: 6. Fluoxetine

7. Paroxetine
8. Mirtazapine
9. Milnacipran

Less good choice for this patient: 10. Nortriptyline

11. Desipramine
12. Bupropion (caution due to seizure disorder history)
13. Protriptyline

Bad choice for this patient, or contraindicated: 14. Citalopram (ineffective based on patient history)

15. Amitriptyline (anticholinergic effects and potential seizure risk)
16. Clomipramine (anticholinergic effects and potential seizure risk)
17. Imipramine (anticholinergic effects and potential seizure risk)
18. Fluvoxamine
19. Isocarboxazid (potential for drug interactions and dietary restrictions)
20. Phenelzine (potential for drug interactions and dietary restrictions)
21. Tranylcypromine (potential for drug interactions and dietary restrictions)

#2

1. Duloxetine
2. Sertraline
3. Escitalopram
4. Fluoxetine
5. Paroxetine
6. Vortioxetine

Good choice for this patient: 7. Venlafaxine

8. Mirtazapine
9. Milnacipran

Less good choice for this patient: 10. Fluvoxamine

11. Desipramine
12. Nortriptyline
13. Protriptyline

Bad choice for this patient, or contraindicated: 14. Bupropion (due to seizure history)

15. Amitriptyline (due to potential seizure risk)
16. Clomipramine (due to potential seizure risk)
17. Imipramine (due to potential seizure risk)
18. Isocarboxazid (due to potential seizure risk)
19. Phenelzine (due to potential seizure risk)
20. Tranylcypromine (due to potential seizure risk)
21. Citalopram (already tried with no reduction in depression symptoms)

#3

1. Sertraline
2. Escitalopram
3. Citalopram
4. Fluoxetine
5. Vortioxetine

Good choice for this patient: 6. Bupropion (considering the history of seizure disorder, use with caution)

7. Duloxetine
8. Paroxetine
9. Fluvoxamine
10. Mirtazapine

Less good choice for this patient: 11. Nortriptyline

12. Desipramine
13. Imipramine
14. Clomipramine
15. Protriptyline

Bad choice for this patient, or contraindicated: 16. Amitriptyline (due to increased seizure risk)

17. Venlafaxine (already tried without success)
18. Isocarboxazid (MAOI with significant dietary restrictions and drug interactions)
19. Phenelzine (MAOI with significant dietary restrictions and drug interactions)
20. Tranylcypromine (MAOI with significant dietary restrictions and drug interactions)
21. Milnacipran (not approved for depression in the United States)

#4

1. Sertraline
2. Vortioxetine

Good choice for this patient: 3. Fluoxetine

4. Citalopram
5. Duloxetine
6. Paroxetine
7. Venlafaxine
8. Mirtazapine

Less good choice for this patient: 9. Bupropion (due to seizure history)

10. Nortriptyline
11. Desipramine
12. Milnacipran
13. Fluvoxamine

Bad choice for this patient, or contraindicated: 14. Amitriptyline (due to seizure history and possible drug interactions)

15. Clomipramine (due to seizure history)
16. Imipramine (due to seizure history)
17. Isocarboxazid (due to possible drug interactions)
18. Phenelzine (due to possible drug interactions)
19. Protriptyline (due to seizure history)
20. Tranylcypromine (due to possible drug interactions)
21. Escitalopram (already tried and ineffective)

#5

1. Best choice for this patient:

- Escitalopram
- Duloxetine
- Fluoxetine
- Citalopram

2. Good choice for this patient:

- Venlafaxine
- Vortioxetine
- Paroxetine
- Mirtazapine

3. Less good choice for this patient:

- Milnacipran
- Fluvoxamine
- Nortriptyline
- Desipramine

4. Bad choice for this patient, or contraindicated:

- Amitriptyline (due to seizure disorder)
- Bupropion (due to seizure disorder)
- Clomipramine (due to seizure disorder)
- Imipramine (due to seizure disorder)
- Isocarboxazid (potential interactions with Ritonavir)
- Phenelzine (potential interactions with Ritonavir)
- Protriptyline (due to seizure disorder)
- Sertraline (ineffective in the past)
- Tranylcypromine (potential interactions with Ritonavir)
